# Neuroimaging markers of patient-reported outcome measures in acute ischemic stroke

**DOI:** 10.1101/2023.12.27.23299829

**Authors:** Lara C. Oliveira, Anna K. Bonkhoff, Robert W. Regenhardt, Kenda Alhadid, Carissa Tuozzo, Mark R. Etherton, Natalia S. Rost, Markus D. Schirmer

## Abstract

**Objectives:** To determine the relationship between patient-reported outcome measures (PROMs) and volumetric imaging markers in acute ischemic stroke (AIS).

**Patients and Methods:** Patients presenting at Massachusetts General Hospital between February 14, 2017 and February 5, 2020 with a confirmed AIS by MRI were eligible and underwent a telephone interview including PROM-10 questionnaires 3-15 months after stroke. White matter hyperintensity (V_WMH_) and brain volumes (V_Brain_) were automatically determined using admission clinical MRI. Stroke lesions were manually segmented and volumes calculated (V_Lesion_). Multivariable and ordinal regression analyses were performed to identify associations between global and PROM-10 subscores with brain volumetrics and clinical variables.

**Results:** Utilizing data from 167 patients (mean age: 64.7; 41.9% female), higher V_WMH_ was associated with worse global physical (β=-0.6), global mental (β=-0.65), physical health (OR=0.68), social satisfaction (OR=0.66), fatigue (OR=0.69) and social activities (OR=0.59) scores. Higher V_Lesion_ was associated with poorer global mental (β=-0.79), mental health (OR=0.68), physical (OR=0.66) and social activities (OR=0.55), and emotional distress (OR=0.68) scores. Higher V_Brain_ was linked to better global mental (β=0.93), global physical (β=0.79), mental health (OR=1.54) and physical activities (OR=1.72) scores.

**Conclusions:** Neuroimaging biomarkers were significantly associated with PROMs, where higher V_WMH_ and V_Lesion_ led to worse outcome, while higher V_Brain_ was protective. The inclusion of neuroimaging analyses and PROMs in routine assessment provides enhanced understanding of post-stroke outcomes.

## INTRODUCTION

Acute ischemic stroke (AIS) is a leading cause of long-term disability^1-3^, with reported 25-74% of the 50 million individuals who have experienced a stroke requiring support from caregivers.^3^ Consequently, understanding the contributors to stroke outcomes is a crucial step to reduce the burden of disability, and could enable healthcare professionals to effectively manage patients’ symptoms and allocate appropriate resources to enhance their recovery.^4^ National Institutes of Health Stroke Scale (NIHSS)^5^ and the modified Rankin Scale (mRS)^6^ scores are valuable and established tools to assess stroke severity and the functional status after stroke. However, both scores fail to capture the full spectrum of symptoms stroke survivors experience and neglect important health domains that are meaningful to the patients, such as mood disorders, fatigue, pain and overall physical activities.^7^ As a result, collecting patient-reported outcome measures (PROMs) is gaining interest, since they allow personalized health assessments of stroke survivors.^8^

Patient-reported outcomes offer a more comprehensive assessment compared to clinician-reported measures.^9^ PROMs have demonstrated reliability and validity across different stroke subtypes and levels of disability, making them suitable for a diverse range of stroke patients.^10^ Moreover, it is crucial to consider patient-reported measures during stroke recovery in order to provide appropriate post-stroke care and a more personalized evaluation of stroke impact with information on physical, social, and cognitive function outcomes. Understanding the determinants for the outcomes that are meaningful to patients is essential for their optimal recovery.

Studies have investigated the determinants of worse stroke outcomes with a particular focus on understanding the association with brain volumetrics.^11-15^ Specifically, white matter hyperintensity (WMH) volume (V_WMH_),^11,16^ brain volume (V_Brain_)^12^ and stroke lesion volume (V_Lesion_)^13^ have been linked to functional outcomes and stroke severity. In recent years, there has been growing interest in quantitatively assessing these phenotypes. Several research groups have developed automated algorithms to quantify WMH volume, although many of them utilize high resolution imaging (Caligiuri et al., 2015; Dadar et al., 2017), which is often not available as part of the clinical assessment of stroke patients. Schirmer et al.^12^ have recently developed and validated a fully automated pipeline that utilizes FLAIR sequences for identifying and segmenting WMH, which also provides a direct assessment of V_Brain_. Although V_Brain_ has also been demonstrated to be an important biomarker associated with IS outcomes, fewer studies consider it when modeling,^12,17,18^ and neuroimaging biomarkers are widely understudied with respect to PROMs.

In this work we utilize the recent developments in neuroimaging analysis for clinical grade MRI data in AIS patients to investigate the relationship between quantitative, volumetric biomarkers of stroke outcome and PROMs. Utilizing a clinical stroke cohort of 167 patients with acute neuroimaging and PROMs data available, we quantify commonly assessed neuroimaging biomarkers and determine their association with patient reported outcome.

## MATERIALS AND METHODS

### Standard protocol approvals, registration, and patient consent

This study is registered under MGH IRB protocol 2019P001189 and uses phenotypic and imaging data collected under MGH IRB protocol 2013P000494. Informed written consent was obtained according to the Declaration of Helsinki from all participating patients or their surrogates.

### Study design, setting, and patient population

We conducted a retrospective analysis using data from a prospective cohort of AIS patients. Patients that were 18 years or older and presented to the Emergency Department at Massachusetts General Hospital between February 2017 and February 2020 with a confirmed diagnosis of AIS using MRI were eligible for inclusion.

### Clinical Assessments

Medical comorbidities and demographic information were extracted for each participant through a review of their medical records at the time of study enrollment. A trained study coordinator assessed pre-stroke disability using the modified Rankin Scale (mRS),^6^ defined as mRS≥1. Telephone interviews assessing PROMs were conducted at 3 to 6 months and/or 6 to 12 months following the stroke, either with the patient or their caregiver, utilizing the PROMIS Global Health (PROMIS GH) questionnaires.^19,20^

The PROMIS short form consists of 10 items (score range 1-5), allowing for the assessment of physical and mental health subscores, where higher scores represent better outcomes.^20^ Each domain within the PROMIS (mental and physical) consists of four subsets of questions, which can then be combined to generate the overall global score. The global mental score is the summation of items 02 (quality of life), 04 (mental health), 05 (social satisfaction), and 10r (emotional distress). The global physical score corresponds to the summation of items 03 (physical health), 06 (physical activities), 07rc (pain), and 08r (fatigue). Moreover, the PROMIS includes assessments for the patient’s social health domain, including assessments for general health (item 01) and social activities (item 09r). T-scores for each of the domains can be derived from the global mental and physical scores, with a score of 50 representing the average score within the US general population (standard deviation (SD): 10).

### Neuroimaging Analysis

Each patient’s FLAIR image underwent automated quantification of V_WMH_ and V_Brain_, utilizing previously published, dedicated pipelines designed for clinical, multi-site neuroimaging data of AIS patients.^21^ In brief, processing included brain extraction (deep-learning based U-net algorithm), intensity normalization using a mean-shift algorithm, and segmentation of WMHs (U-net based deep-learning algorithm).^21^ Details on the specific algorithms and their validations can be found elsewhere.^12,21^

Stroke lesions were manually delineated by a vascular neurologist (LCO) using the acute diffusion weighted imaging data (b=1000) and 3D Slicer (https://www.slicer.org/).^22^ Combined, these steps resulted in 3 segmentation masks per patient, through which V_Brain_, V_WMH_, and V_Lesion_ were calculated, given by the voxel count within the mask multiplied by the voxel volume in cm^3^.

Furthermore, data and segmentation quality was assessed by a trained vascular neurologist (LCO) and visually inspected to ensure accurate delineation and patients whose segmentation masks did not pass the quality assessment were subsequently excluded from the analysis.

### Statistical analysis and model description

Summary statistics of the cohort characteristics were calculated. Normal-distributed variables were summarized as mean and standard deviation (SD), and non-normal distributed variables as median and interquartile range (IQR). As appropriate, ANOVA, for normal-distributed variables, Kruskal-Wallis, for non-normal distributed variables, and χ^2^ tests for categorical variables, were used to evaluate differences between included and excluded participants in the analysis. Moreover, continuous variables were z-transformed and extreme values were identified as outliers if data points with z-scores ≥3. These patients were subsequently removed before modeling. Subsequently, lesion loads WMHL and LesionL were calculated as V_i_/V_Brain_ for WMH and stroke lesion, respectively.

Regression models were used to investigate the association between PROMs with brain volumetrics (V_Brain_, V_WMH_, and V_Lesion_), while accounting for age, sex and common cardiovascular risk factors (hypertension and diabetes). Regression models were used, given by

Outcome ~ Age + pre-stroke disability + DM2 + HTN + Sex + V_Brain_ + logit(LesionL) + logit(WMHL), where Outcome represents either the mental or physical T-scores as dependent variables (multivariable linear regression) or individual items of the PROMIS questionnaire (ordinal regression). Independent factors include diabetes (DM2), hypertensive status (HTN), female sex (Sex), brain volume (V_Brain_), acute lesion load (LesionL), and WMH load (WMHL).

Assumptions of each regression were examined. Multivariable regression model assumptions include normality of residuals, linearity, and homoscedasticity through diagnostic plots, and collinearity among the predictor variables using variance inflation factor (VIF) values, with conservative threshold of VIF < 2 indicating no significant collinearity. For ordinal regressions, the proportional odds assumption was assessed using the Brant test.^23^

Since outcomes were collected at one of two time points post-stroke, we included the time point at which PROMs were obtained as a nuisance factor in all models. All statistical analyses were conducted using the computing environment R Version 4.1.1 (R Foundation for Statistical Computing, Vienna, Austria). Significance was set at *P<0*.*05*.

### Data availability statement

The authors have agreed to make the data, analysis methods, and research materials available to other researchers for the purpose of replicating the results. This will be done with the explicit permission of the local institutional review board, ensuring compliance with data sharing regulations and protection of patient privacy.

## RESULTS

A total of 167 patients with available PROMIS data at follow-up were utilized in this study. The cohort was on average 64.7 +/-12.4 years old, and 41.9% were female. Of the 167 patients, 17 were excluded from analysis, of which 4 patients did not meet the quality criteria for the segmentation masks, and 13 patients were identified as outliers. Compared to the excluded population, the included participants were older (65.3 vs. 58.8, *P=*.*04*) and had higher rates of hypertension (78.4% vs. 50.0%, *P=*.*03*). Additionally, they had a greater burden of WMH (3.1 cc vs. 1.4 cc, *P=*.*03*). The cohort characteristics are described in Table 1.

**Table 1.** Characteristics of the study cohort. Patients were excluded if they did not meet quality control standards for the volumetric segmentation (N=4) or if they were identified as outliers for the analysis (N=13).

| Variables | Total<br>N = 167 | Included<br>N = 150 | Excluded<br>N = 17 | P value |
| --- | --- | --- | --- | --- |
| Age (mean (SD)) | 64.7 (12.4) | 65.3 (12.1) | 58.8 (13.6) | **.04** |
| Female sex, n (%) | 70 (41.9) | 63 (42.0) | 7 (41.2) | 1.0 |
| **Medical History, n (%)** |  |  |  |  |
| Hypertension | 124 (75.6) | 116 (78.4) | 8 (50.0) | **.03** |
| Diabetes | 41 (24.8) | 40 (27.0) | 1 (5.9) | .11 |
| Pre-stroke disability<br>(mRS<sub>pre</sub> > 0) | 39 (23.6) | 36 (24.3) | 3 (17.6) | .76 |
| **Image Data, median [IQR]** |  |  |  |  |
| V<sub>WMH</sub> cm<sup>3</sup> | 2.6 [1.3, 6.8] | 3.1 [1.4, 7.0] | 1.4 [0.5, 2.1] | **.03** |
| V<sub>Lesion</sub> cm<sup>3</sup> | 2.2 [0.6, 10.7] | 2.1 [0.5, 8.5] | 4.4 [1.0, 16.7] | .21 |
| V<sub>Brain</sub> cm<sup>3</sup> | 1312.1 [1192.4, 1414.0] | 1307.3 [1188.9, 1410.6] | 1367.6 [1249.1, 1416.9] | .39 |
| **Outcomes** |  |  |  |  |
| T Physical (mean (SD)) | 47.4 (9.4) | 47.5 (9.3) | 46.5 (10.9) | .69 |
| T Mental (mean (SD)) | 48.2 (8.9) | 48.2 (8.7) | 48.4 (10.9) | .94 |

mRS, modified Rankin Scale; V<sub>Brain</sub>, brain volume; V<sub>Lesion</sub>, stroke lesion volume; and V<sub>WMH</sub>, white matter hyperintensities volume. P values < 0.05 are in bold.

The results of the regression models are summarized in Table 2 and Figure 1. All models fulfilled the assumptions. Increasing age was associated with better physical (β=3.09 [1.26–4.92], *P=*.*001*) and mental (β=2.90 [1.16–4.63], *P=*.*001*) T-scores, while pre-stroke disability was associated with worse physical T-scores (β=-5.38 [-8.86–-1.90], *P<*.*01*). In relation to brain volumetrics, larger brain volume was associated with better physical (β=2.47 [0.54–4.40], *P=*.*01*) and mental (β=2.33 [0.49–4.16], *P=*.*02*), larger volumes of stroke lesions were associated with worse mental (β=-2.02 [-3.40– -0.64], *P<*.*01*), and higher WMH burden was associated with both lower physical (β=-1.90 [-3.51–-0.29], *P=*.*02*) and mental (β=-1.64 [-3.16–-0.11], *P=*.*03*) T-scores (Table 2 and Figure 1A).

**Table 2.** Estimates and Odds Ratios of regression models. Columns contain the PROMs T scores and subscores as the dependent variables of our statistical model, while rows contain the independent variables.

|  | T Physical | T Mental | Global 01 | Global 02 | Global 03 | Global 04 | Global 05 | Global 06 | Global 07rc | Global 08r | Global 09r | Global 10r |
| --- | --- | --- | --- | --- | --- | --- | --- | --- | --- | --- | --- | --- |
| | $\beta$<br>(CI)<br>P value | $\beta$<br>(CI)<br>P value | OR<br>(CI)<br>P value | OR<br>(CI)<br>P value | OR<br>(CI)<br>P value | OR<br>(CI)<br>P value | OR<br>(CI)<br>P value | OR<br>(CI)<br>P value | OR<br>(CI)<br>P value | OR<br>(CI)<br>P value | OR<br>(CI)<br>P value | OR<br>(CI)<br>P value |
| Age | 3.09<br>1.26 , 4.92<br>**.001** | 2.90<br>1.16 , 4.63<br>**.001** | 2.33<br>1.55 , 3.54<br>**<0.001** | 1.60<br>1.09 , 2.36<br>**0.02** | 2.06<br>1.37 , 3.13<br>**0.001** | 1.62<br>1.10 , 2.42<br>**0.02** | 1.90<br>1.29 , 2.83<br>**<0.01** | 2.17<br>1.44 , 3.32<br>**<0.001** | 1.51<br>1.00 , 2.28<br>0.05 | 1.32<br>0.88 , 1.97<br>0.18 | 1.76<br>1.20 , 2.62<br>**<0.01** | 1.73<br>1.16 , 2.62<br>**<0.01** |
| Sex(F) | -0.66<br>-4.31 , 3.00<br>.72 | 1.13<br>-2.34 , 4.60<br>.52 | 0.57<br>0.25 , 1.28<br>.18 | 0.82<br>0.36 , 1.81<br>0.62 | 0.59<br>0.25 , 1.36<br>0.22 | 1.86<br>0.84 , 4.17<br>0.13 | 0.98<br>0.44 , 2.16<br>0.95 | 0.69<br>0.29 , 1.61<br>0.39 | 0.82<br>0.37 , 1.80<br>0.62 | 0.65<br>0.28 , 1.47<br>0.30 | 0.86<br>0.38 , 1.92<br>0.72 | 0.86<br>0.38 , 1.91<br>0.71 |
| mRS<sub>pre</sub>><br>0 | -5.38<br>-8.86 , -1.90<br>**<0.01** | -0.28<br>-3.58 , 3.02<br>.87 | 0.65<br>0.31 , 1.35<br>0.25 | 0.50<br>0.23 , 1.06<br>0.07 | 0.36<br>0.17 , 0.78<br>**0.01** | 1.24<br>0.59 , 2.62<br>0.57 | 1.09<br>0.53 , 2.26<br>0.82 | 0.35<br>0.16 , 0.74<br>**<0.01** | 0.37<br>0.17 , 0.79<br>**0.01** | 0.69<br>0.33 , 1.44<br>0.33 | 0.94<br>0.44 , 1.98<br>0.86 | 1.61<br>0.77 , 3.39<br>0.21 |
| HTN | -0.73<br>-4.38 , 2.91<br>.69 | -3.18<br>-6.63 , 0.28<br>.07 | 0.42<br>0.19 , 0.92<br>**0.03** | 0.47<br>0.22 , 1.02<br>0.06 | 0.69<br>0.31 , 1.52<br>0.36 | 0.72<br>0.32 , 1.63<br>0.44 | 0.53<br>0.24 , 1.14<br>0.11 | 0.96<br>0.40 , 2.27<br>**0.93** | 1.30<br>0.61 , 2.77<br>0.50 | 0.80<br>0.36 , 1.78<br>0.59 | 0.72<br>0.32 , 1.61<br>0.43 | 0.53<br>0.24 , 1.15<br>0.11 |
| DM2 | -1.17<br>-4.60 , 2.26<br>.50 | -1.47<br>-4.73 , 1.78<br>.37 | 0.54<br>0.25 , 1.16<br>0.12 | 0.49<br>0.23 , 1.03<br>0.06 | 0.69<br>0.32 , 1.47<br>0.34 | 0.65<br>0.32 , 1.33<br>0.25 | 0.87<br>0.42 , 1.79<br>0.71 | 0.46<br>0.22 , 0.97<br>**0.04** | 0.81<br>0.39 , 1.69<br>0.57 | 0.97<br>0.46 , 2.02<br>0.94 | 0.46<br>0.22 , 0.97<br>**0.04** | 0.69<br>0.33 , 1.46<br>0.34 |
| V<sub>Brain</sub> | 2.47<br>0.54 , 4.40<br>**.01** | 2.33<br>0.49 , 4.16<br>**.01** | 1.37<br>0.89 , 2.11<br>0.16 | 1.30<br>0.87 , 1.96<br>0.21 | 1.30<br>0.84 , 2.01<br>0.25 | 1.54<br>1.02 , 2.35<br>**0.04** | 1.37<br>0.91 , 2.09<br>0.14 | 1.72<br>1.09 , 2.76<br>**0.02** | 1.40<br>0.92 , 2.15<br>0.12 | 1.17<br>0.75 , 1.80<br>0.48 | 1.27<br>0.83 , 1.95<br>0.27 | 1.32<br>0.86 , 2.03<br>0.20 |
| WMHL | -1.90<br>-3.51 , -0.29<br>**.02** | -1.64<br>-3.16 , -0.11<br>**.04** | 0.81<br>0.57 , 1.14<br>0.23 | 0.79<br>0.56 , 1.10<br>0.17 | 0.68<br>0.47 , 0.98<br>**0.04** | 0.76<br>0.54 , 1.07<br>0.12 | 0.66<br>0.47 , 0.92<br>**0.02** | 0.70<br>0.48 , 1.01<br>0.06 | 1.07<br>0.75 , 1.52<br>0.71 | 0.69<br>0.49 , 0.98<br>**0.04** | 0.59<br>0.41 , 0.83<br>**<0.01** | 0.84<br>0.59 , 1.19<br>0.33 |
| LesionL | -1.24<br>-2.70 , 0.21<br>.09 | -2.02<br>-3.40 , -0.64<br>**<0.01** | 0.99<br>0.72 , 1.35<br>0.93 | 0.76<br>0.56 , 1.04<br>0.09 | 0.78<br>0.57 , 1.08<br>0.14 | 0.68<br>0.50 , 0.93<br>**0.02** | 0.80<br>0.58 , 1.09<br>0.16 | 0.66<br>0.46 , 0.93<br>**0.02** | 0.94<br>0.69 , 1.28<br>0.70 | 0.88<br>0.64 , 1.20<br>0.41 | 0.55<br>0.40 , 0.76<br>**<0.001** | 0.68<br>0.50 , 0.93<br>**0.02** |

$\beta$ , the coefficients for the linear regression models; DM2, diabetes; HTN, hypertension; LesionL, stroke lesion load; mRS<sub>pre</sub>, modified Rankin Scale; OR, odds ratios for ordinal regression models; Sex(F), female sex; V<sub>Brain</sub>, brain volume; and WMHL, white matter hyperintensities lesion load. P values < 0.05 are in bold.

**Figure 1.**
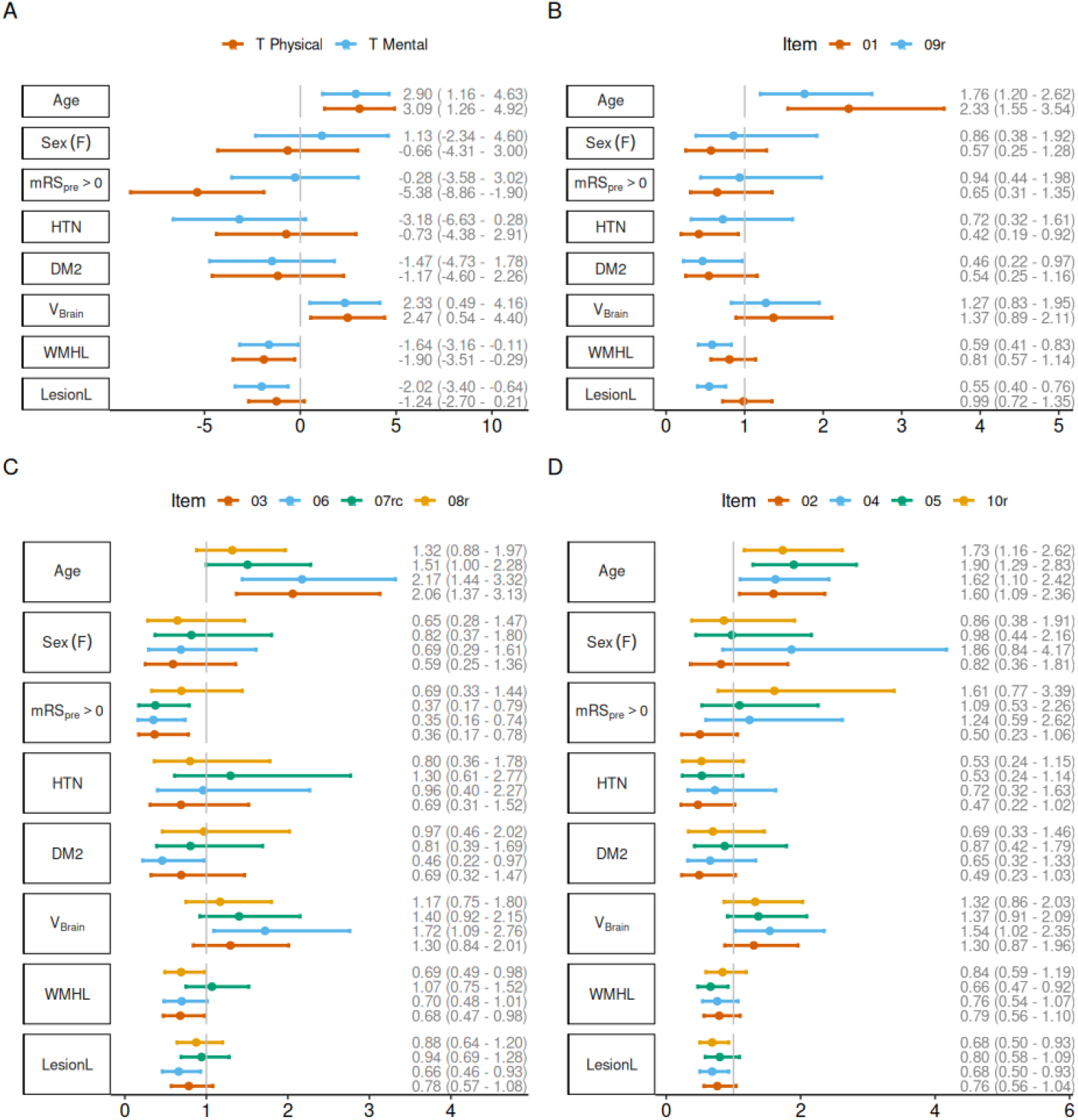
Regression models overview. A) Multivariable linear regression for T Mental and T Physical scores. B) Ordinal regression models for general health and social activities. C) Ordinal regression models for physical health subitems. D) Ordinal regression models for mental health subitems. *DM2, diabetes; HTN, hypertension; Item 01, general health; Item 02, quality of life; Item 03, physical health; Item 04, mental health; Item 05, social satisfaction; Item 06, physical activities; Item 07rc, pain; Item 08r, fatigue; Item 09r, social activities; Item 10r, emotional distress; LesionL*, *stroke lesion load; mRS*_*pre*_,, *pre-stroke mRS; V*_*Brain*,_ *brain volume; WMHL white matter hyperintensity load*.

In the physical domain (Figure 1C, Table 2), advanced age was linked to better physical health (Item 03 - OR 2.06 [1.37–3.13], *P=*.*001*) and physical activities (Item 06 - OR 2.17 [1.44–3.32], *P<*.*001*). Pre-stroke disability was independently associated with worse patient-reported physical outcomes (Item 03: OR 0.36 [0.17, 0.78–], *P=*.*01*, Item 06: OR 0.35 [0.16–0.74], *P<*.*01*, and Item 07rc: OR 0.37 [0.17–0.79], *P=*.*01*). Additionally, the presence of diabetes was an independent predictor of poorer physical activities (Item 06: OR 0.46 [0.22–0.97], *P=*.*04*). Furthermore, brain volume was linked to better (Item 06 - OR 1.72 [1.09–2.76], *P=*.*02*), while stroke lesion volume was associated with worse patient-reported physical activities scores (Item 06 - OR 0.66 [0.46–0.93], *P=*.*02*). Moreover, higher burden of WMH was a predictor of worse scores in physical health (Item 03 - OR 0.68 [0.47–, 0.98], *P=*.*04*) and fatigue (Item 08r - OR 0.69 [0.49–0.98], *P=*.*04*).

In the mental domain (Figure 1D, Table 2), increased age was associated with better scores for all mental PROMIS subcomponents, including quality of life (Item 02 - OR 1.60 [1.09–2.36], *P=*.*02*), mental health (Item 04 - OR 1.62 [1.10–2.42], *P=*.*02*), social satisfaction (Item 05 - OR 1.90 [1.29–2.83], *P<*.*01*), and emotional problems (Item 10r - OR 1.73 [1.16–2.62], *P<*.*01*). Moreover, larger brain volumes were found to be associated with better mental health (Item 04 - OR 1.54 [1.02–2.35], *P=*.*04*). Larger lesion volumes were predictors for worse mental health (Item 04 - OR 0.68 [0.50–0.93], *P=*.*02*) and emotional problems (Item 10r - OR 0.68 [0.50–0.93], *P=*.*02*), and a higher burden of WMH was linked to worse social satisfaction (Item 05 - OR 0.66 [0.47–0.92], *P=*.*02*).

Increasing age was linked to better patient-reported general health (Item 01 - OR 2.33 [1.55–3.54], *P<*.*001*) and social activities (Item 09r - OR 1.76 [1.20–2.62], *P<*.*01*). Additionally, hypertension was associated with lower general health (Item 1) scores (OR 0.42 [0.19–0.92], *P=*.*03*), and diabetes with worse social activities (Item 09r - OR 0.46 [0.22–0.97], *P=*.*04*). Similarly, higher lesion volumes and WMH burden were associated with lower social activities (Item 09r - OR 0.55 [0.40–0.76], *P<*.*001* and OR 0.59 [0.41–0.83], *P<*.*01*, respectively).

## DISCUSSION

In this study, we evaluated associations between neuroimaging biomarkers and PROMs in AIS patients. Our findings underscore the importance of the relationship between patient-reported outcome measures and brain volumetrics, specifically with brain, stroke lesion, and WMH volumes.

Our study reveals that higher WMH burden is associated with worse T scores for mental and physical health. Specifically, sub-scores related to physical health, social satisfaction, fatigue, and social activities are negatively affected by WMH volume. Previous research has investigated the association between WMH burden and worse functional outcomes following stroke, primarily assessed using the mRS,^11,16,24,25^ demonstrating that increased WMH burden negatively affects stroke outcomes.^24,16^. Similarly, our study shows that WMH burden is linked with worse physical health (Item 03), which closely reflects outcome measured by mRS.

Furthermore, our results support that V_Brain_ serves as a protective mechanism, leading to better T scores for mental and physical health. This is also reflected in Items 04 (mental health) and 06 (physical activities). Our results align with previous research, which has demonstrated an association between greater brain volume and lower mRS, i.e. better outcomes.^12^ These findings may reflect the brain’s ability to compensate for injuries and disease burden, which has previously been explored in stroke patients in the form of effective reserve.^26^

Additionally, we report on the unique association between stroke lesion volume and PROMs. Our results indicate that larger lesion volumes are associated with poorer patient-reported mental outcomes, indicated by the T score and mental subscores for mental health, physical activities, social activities, and emotional problems. While it is known that larger lesion volumes are linked to functional disability after stroke,^13^ our findings highlight the relationship between V_Lesion_ and PROMs beyond commonly assessed motor functions. In general, mRS is considered to have a strong bias toward physical disability, as it primarily assesses motor function.^6^ However, it is important to recognize that a disabling mRS may impact other non-physical factors that are vital to an individual’s self-care and overall well-being.^6^ These factors encompass social abilities, social functioning, and mood disturbances, all of which contribute to the perception of disability.^27^ Our prior research, examining stroke outcomes in both sexes, revealed that a mRS score of 2 or higher was associated with poorer T mental scores in both sexes.^28^ However, additional work is needed to enhance our understanding of the complimentary information between mRS and PROMs.

Our results also unveil that increasing age is associated with better patient-reported outcomes, contrary to the common belief that older patients are to expect worse outcomes. Previous studies have also demonstrated a positive association between advancing age and improved patient-reported outcomes following orthopedic and oncological surgeries.^29-31^ These findings imply that in younger age groups, there may be other significant factors related to functional requirements that need to be considered. Additionally, unrealistic expectations of recovery can contribute to poorer self-reported outcomes in younger patients.^30^

This study has some limitations. Firstly, the patient evaluation after discharge was conducted solely through phone interviews, which may have introduced a selection bias by including only patients and/or caregivers who were willing and able to participate in phone conversations. Moreover, some patients were not available at the first time window of the phone interview. As a result, a second time window for reassessment was implemented. However, in order to reduce the effect of different time windows, we specifically included the time point at which PROMs were obtained as a nuisance factor in our models. Secondly, our study was carried out at a single comprehensive stroke center, which could potentially limit the generalizability of our findings to a broader population. However, it is important to note that our stroke center has a wide geographic catchment area, including patients from the largest telestroke network in New England. While the standardized protocols and rigorous methodology employed increase the reliability and validity of our results, we acknowledge the need for further studies, involving multiple centers to confirm the generalizability of our findings. Lastly, it is worth noting that our study population consists of patients with relatively small ischemic strokes (median V_Lesion_ of 2cm^3^), which is common in other imaging studies of acute ischemic stroke.^33,33^ Future studies that aim to recruit a broader population in terms of lesion volume/stroke severity, such as the DISCOVERY study,^34^ are required to ensure the applicability of our findings across a wider range of stroke cases.

Strengths of our study include the utilization of state-of-the-art clinical neuroimaging analysis methodologies, allowing us to delve deeper into the associations with post-stroke outcomes at the time of admission to the hospital. Importantly, to the best of our knowledge, this study represents the first investigation into the association between neuroimaging markers and patient-reported outcomes in AIS. Additionally, our cohort size of 150 patients allowed us to include the most common risk factors associated with other functional outcome measures into our models. Finally, our study provides an important first step for comprehending the factors that contribute to PROMs, particularly the features from initial neuroimaging at stroke presentation, uncovering detrimental and protective determinants, expanding our knowledge of stroke recovery and outcomes.

In conclusion, our study shows that WMH, brain, and stroke lesion volume are important biomarkers for PROMs that can be readily assessed using clinical neuroimaging data in the emergency department. The inclusion of neuroimaging analyses and patient-reported outcome measures has the potential to provide enhanced understanding of post-stroke recovery.

